# Principles and performance of wastewater concentration methods for environmental surveillance of viruses: a systematic review and meta-analysis

**DOI:** 10.64898/2026.03.19.26348821

**Authors:** Joyce Odeke Akello, Ben Bellekom, Alexander G. Shaw, Nicholas C Grassly

## Abstract

Methods to concentrate wastewater samples are essential for sensitive environmental surveillance of infectious diseases. We defined six main principles used to concentrate viral pathogens in wastewater and performed a systematic review and meta-analysis of their performance. PubMed and Web of Science were searched on 31 January 2025 using terms wastewater, sewage, concentration methods and wastewater surveillance. We included all studies comparing ≥2 concentration methods for virus detection. Our search identified 49 eligible studies published since 2013 across seven continents. We ranked the performance of evaluated methods in each study and generated an overall performance metric for each method principle by virus group (enveloped vs. non-enveloped) using Plackett-Luce analysis. Precipitation and filtration methods were the most studied, while magnetic bead-based and centrifugation were least studied. Magnetic bead-based methods were more effective for concentrating enveloped viruses (63% of pairwise comparisons), whereas flocculation performed better for non-enveloped viruses (60%). However, no single method strongly dominated and method rankings were variable between studies. This study provides evidence-based guidance for selecting wastewater concentration methods to support environmental surveillance of viral pathogens.

## Main

Wastewater and environmental surveillance (WES) has been used for decades to monitor pathogens such as poliovirus, cholera, and typhoid in the population [1-3]. Its importance became particularly evident during the COVID-19 pandemic, when implementation of wastewater surveillance for SARS-CoV-2 demonstrated its value as an early warning system capable of detecting rising infection trends, predicting epidemic peaks, and identifying imported or novel variants of concern (VOCs) [4-6]. These developments have driven a renewed interest in WES as a powerful tool for infectious disease surveillance, marking a renaissance in the field [7-11]. Critically important to WES is the wastewater concentration method used to recover pathogens present at low abundance for downstream detection. Viable pathogen particles and/or their genomes such as poliovirus, *Salmonella Typhi*, SARS-CoV-2 are shed in bodily excreta including faeces and urine which are subsequently disposed of in wastewater and sewage systems [12-15]. Wastewater comprises a heterogeneous mixture of domestic, institutional, industrial, and surface runoff inputs, whereas sewage refers specifically to human excreta and associated waste streams, as defined by ISO 8099-1 (ISO, 2018). Unlike sewage, wastewater does not necessarily contain faecal material, further complicating pathogen detection.

Identifying the diversity of pathogens in wastewater is substantially more challenging than in clinical samples, primarily due to the extremely low concentrations at which they are present. This typically results from dilution of the wastewater by rainfall, agricultural run-off, industrial or commercial discharge, and residential wastewater including that from bathing and cooking [16]. To ensure recovery of the low concentration pathogens, concentration of large volumes of wastewater into a much smaller volume is usually required for pathogen detection assays such as PCR, cell culture, or a combination of both. Therefore, a concentration step is critical to achieving reliable pathogen detection signals and avoiding false-negative results. In addition to dilution, the wastewater matrix presents significant analytical challenges. Wastewater contains diverse microbial communities, a complex mix of organic and inorganic matter, chemical contaminants, and PCR inhibitors such as humic acid or fumaric acid derived from soil [17-19]. These components can interfere with pathogen concentration methods by adsorbing to concentration materials, clogging filters, or inhibiting downstream detection approaches, thereby reducing overall recovery efficiency and assay sensitivity [20].

Wastewater concentration methods refers to the techniques used to increase the concentration of pathogens present in the sample, enhancing detection and characterisation. Many wastewater concentration methods have been developed and refined to address these challenges. These methods can be categorised based on their underlying principles, including i) centrifugation, ii) filtration, iii) flocculation, iv) magnetic bead-based approaches, v) precipitation, and vi) ultrafiltration (Fig. 1) [21-23]. Centrifugation-based approaches rely on density-driven sedimentation under centrifugal force to pellet larger and denser particles, including viruses. Filtration-based methods exploit electrostatic interactions between viral particles and charged filter surfaces, enabling adsorption to electropositive filters or to electronegative filters via salt bridging mediated by multivalent cations, followed by elution and, where necessary, reconcentration [24-26]. Flocculation-based methods induce chemically mediated aggregation of viral particles into flocs that can be separated from the wastewater matrix [27]. Magnetic bead–based approaches use functionalised beads to selectively capture viral targets via electrostatic, affinity, glycan-mediated, or antibody and receptor-based interactions, enabling magnetic separation from the wastewater matrix [28, 29]. Precipitation-based methods modify sample physicochemical properties typically through the addition of salts or polymers and pH adjustment to reduce solubility and induce aggregation of pathogens and other macromolecules, which are then recovered by centrifugation or filtration [30, 31]. Finally, ultrafiltration concentrates pathogens through size-based retention on semi-permeable membranes, allowing recovery of microorganisms larger than the membrane cut-off, followed by elution and, where required, secondary reconcentration for downstream analysis.

**Fig. 1:**
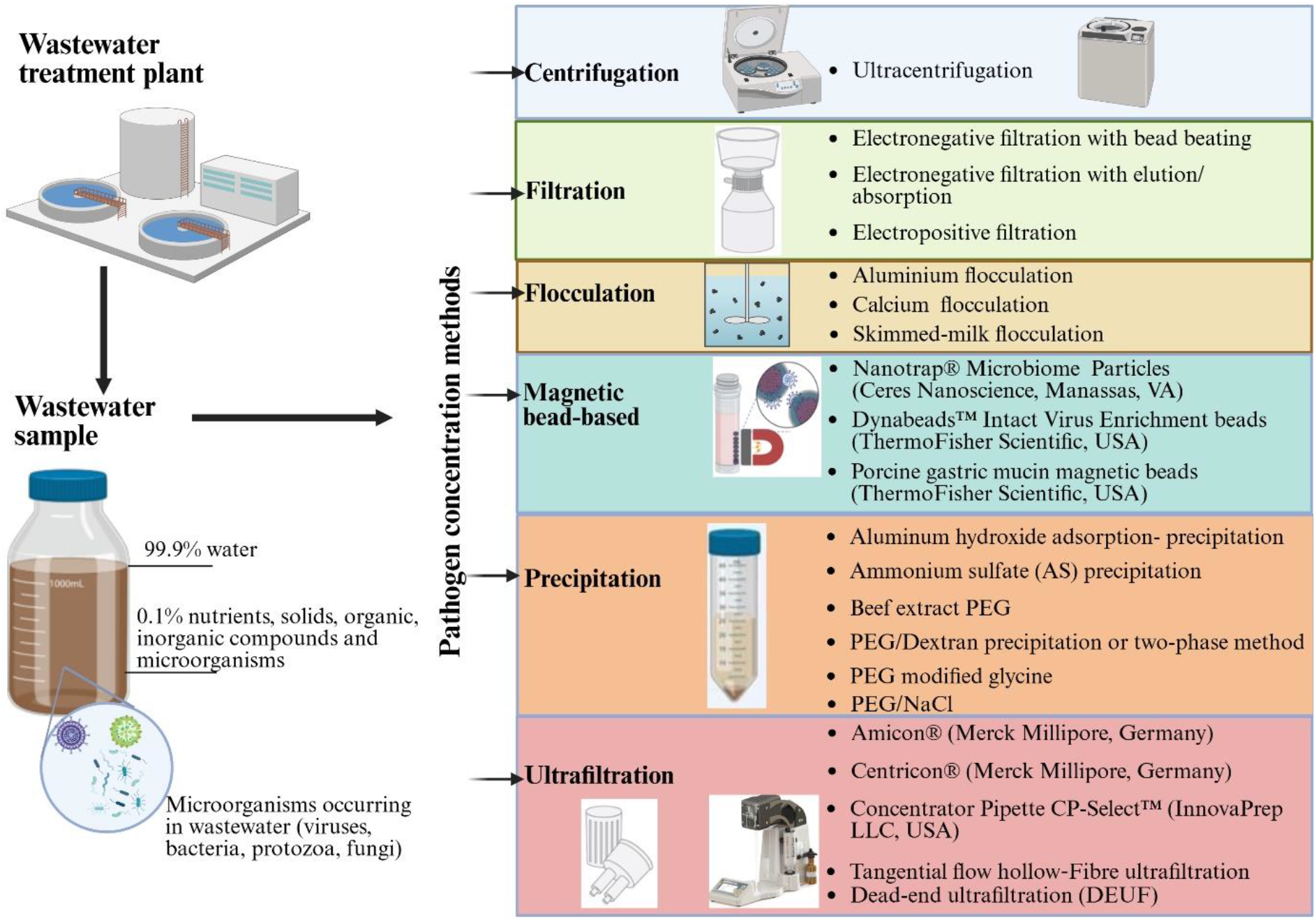
Wastewater concentration methods can be classified into six broad groups, namely, (1) centrifugation, (2) filtration, (3) flocculation, (4) magnetic bead-based, (5) precipitation, and (6) ultrafiltration. Example of the method variants falling within each concentration method group and products are listed. Created in BioRender. https://BioRender.com/ygyhfcr.

Wastewater pathogen concentration performance is influenced by three key factors: sample type, sample composition, and sample volume (Fig. S1). Different wastewater sources, including municipal wastewater, industrial effluent, agricultural runoff, and sludge vary substantially in their physical, chemical, and biological characteristics, which can affect virus stability, partitioning, and pathogen recovery efficiency [32, 33]. In addition, parameters such as turbidity, total solids, organic load, pH, and microbial biomass, together with the volume of sample processed, can influence both concentration performance and downstream detection sensitivity [34, 35]. The applicability of wastewater concentration methods depends strongly on wastewater sample type. Raw influent wastewater, commonly used for viral surveillance, typically contains dilute pathogens and favours precipitation-based, filtration, and ultrafiltration methods. Primary sludge is enriched in pathogen-associated solids and is best suited to centrifugation-based approaches. Magnetic bead–based methods have been successfully applied across both wastewater sample types. However, the high PCR inhibitor content in sludge necessitates careful optimisation of concentration methods to ensure robust pathogen detection. Detailed narrative reviews of each concentration method, including methodological variants and performance considerations, are provided in the supplementary information (Supplementary Appendix).

While methodological advances have aimed to improve pathogen recovery efficiencies, increase input sample volumes, reduce processing time and cost, and minimise co-concentration of inhibitors, all approaches impose physical or chemical stresses that can affect pathogen integrity and detection by molecular or other methods. Moreover, each method has inherent limitations, including variable recovery efficiencies, non-homogeneous enrichment of pathogens by group or species, and methodological biases arising from operational parameters [36-38]. Importantly, most concentration methods target only a single fraction of the wastewater matrix, such as either the solid or supernatant fraction, limiting their suitability for simultaneous detection of diverse pathogens with differing physicochemical properties [38, 39].

Despite extensive method development, no wastewater concentration method has yet achieved universal applicability across the wide range of pathogens and wastewater characteristics encountered in practice. To assess the occurrence of infectious disease-related pathogens in wastewater, it is well recognised that optimal methods should be rapid, simple, consistent, sensitive, and cost-effective to support routine surveillance [40, 41]. However, the diversity of wastewater matrices and pathogen characteristics continues to hinder the establishment of a single standardised method capable of reliably recovering all relevant pathogens.

Although several reviews have discussed the importance of WES and the general challenges associated with pathogen detection in wastewater, a comprehensive systematic evaluation of wastewater concentration methods and their performance in pathogen recovery is lacking. This systematic review and meta-analysis addresses this gap by critically examining wastewater concentration methods as a key determinant of effective environmental pathogen surveillance. We synthesise evidence on the performance of different concentration methods, with a particular focus on viral pathogen recovery, and discuss how method selection influences sensitivity, reliability, and applicability for integrated public health surveillance. Ultimately, this review aims to provide practical guidance for researchers and practitioners seeking robust and evidence-based wastewater concentration strategies to support infectious disease monitoring.

## Results

### Study characteristics

A total of 229 articles were identified through the database search. After removal of duplicate records (n = 16), 213 unique articles remained for screening. Of these, 128 records were excluded based on title and abstract screening, and a further 36 full-text articles were excluded for not meeting the inclusion criteria or meeting one or more exclusion criteria. Ultimately, 49 studies were included in the meta-analysis from the 85 full-text articles assessed for eligibility. The study selection process is illustrated in Fig. 2.

**Fig. 2:**
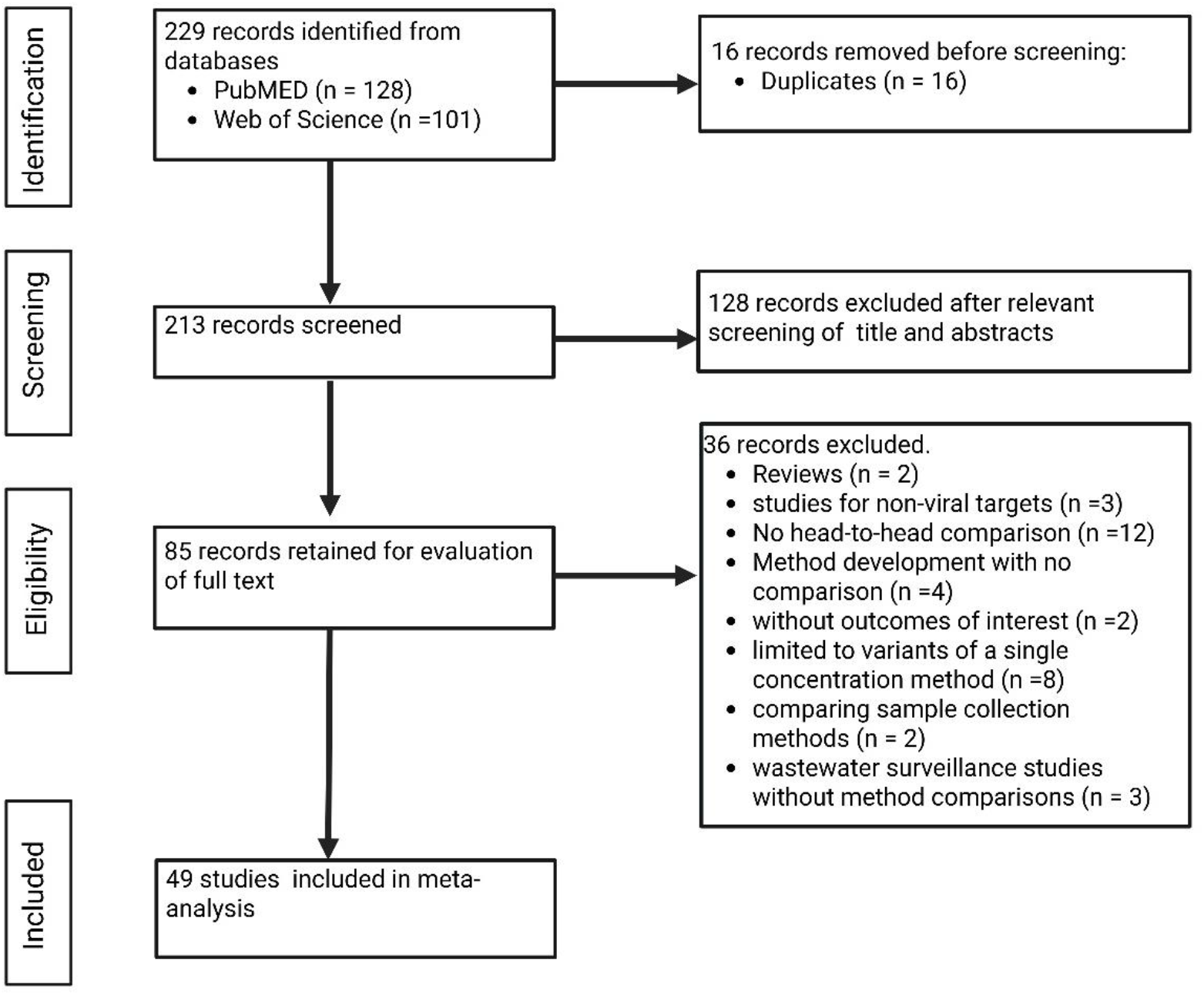
Study selection process following the PRISMA reporting guidelines for systematic review.

The included studies evaluated wastewater concentration methods representing the six major concentration mechanisms: centrifugation (10 studies), filtration (29 studies), flocculation (15 studies), magnetic bead–based methods (9 studies), precipitation (33 studies), and ultrafiltration (19 studies). Several studies evaluated more than one concentration approach. The 49 studies were published between the year 2013 and January 2025. Of these, 45 studies evaluated enveloped viruses, while 20 studies evaluated non-enveloped viruses. The studies were geographically diverse, with research conducted in North America (n = 19), South America (n = 5), Europe (n = 7), Asia (n = 9), Africa (n = 3), Australia (n = 5), and the Caribbean (n = 1) (Fig. 3). Overall, the body of evidence was dominated by studies conducted in high-income countries, with fewer studies originating from low- and middle-income settings.

**Fig. 3:**
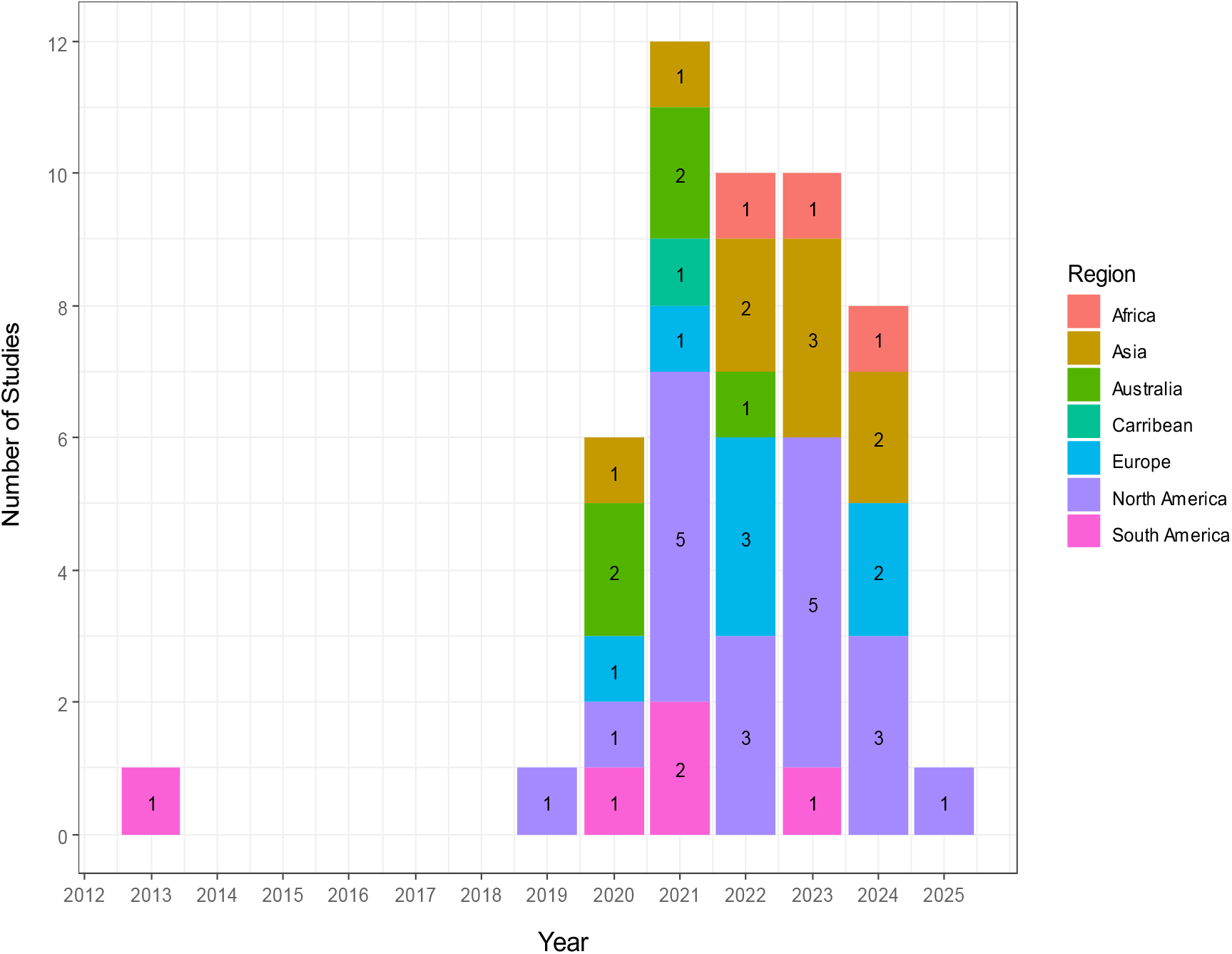
Number of studies included in the systematic review by year of publication. Colours indicate the seven geographical regions where the research studies were conducted. Majority of the studies were from North America. There was an increase in the number of studies evaluating different wastewater concentration methods from the year 2020 onward following the COVID-19 pandemic, with the highest number of studies (n = 12) conducted in the year 2021.

Six pathogen detection techniques were reported across the included studies: quantitative PCR (qPCR), digital PCR (dPCR) and droplet digital PCR (ddPCR), plaque assay, cell culture, and next-generation sequencing (NGS) based approaches (Fig. 4A). qPCR was the most used detection method, employed in 44 out of 49 studies, either alone or in combination with other techniques. Digital PCR methods were used in nine studies, including two using dPCR alone, three using ddPCR alone, and four combining digital PCR with qPCR. Plaque assays were used in one study, cell culture combined with qPCR in one study, metagenomic sequencing in one study, and targeted NGS approaches in one study alone and in two studies in combination with qPCR. SARS-CoV-2 was the most frequently detected and tested pathogen from 2020 onward during the COVID-19 pandemic, reflecting heightened interest in implementing wastewater surveillance for SARS-CoV-2 during this period. Prior to the pandemic, methodological comparisons primarily focused on enteric viruses, including adenovirus, norovirus, rotavirus, and hepatitis A (Fig. 4B).

**Fig. 4:**
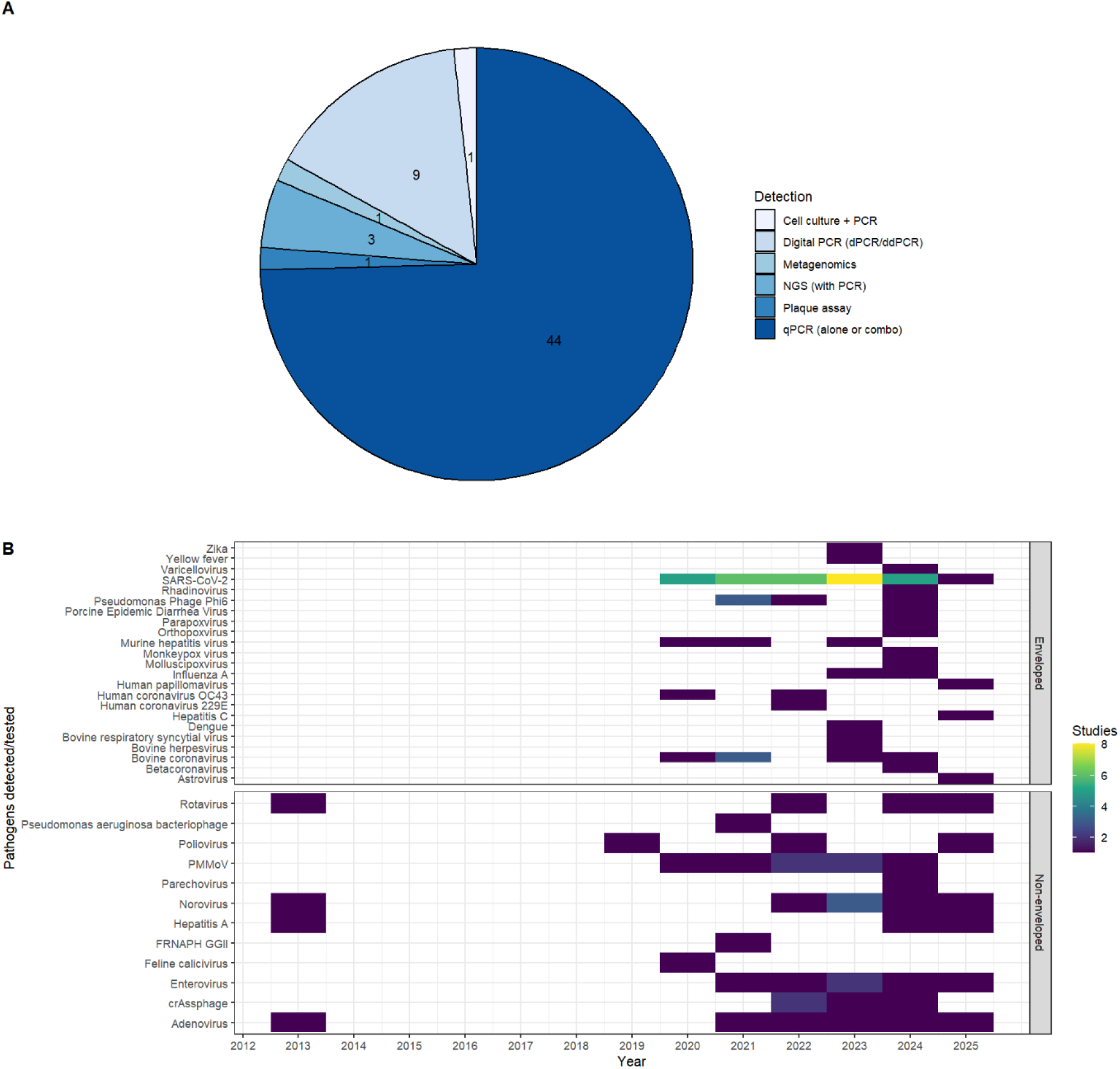
(A) Detection methods used across studies by frequency. (B) Pathogens detected / tested across studies by year of publication. qPCR was the most used method for pathogen detection (A) and there was an increase in the diversity of viral pathogens being detected and tested between the year 2020 and 2025 (B).

### Concentration methods meta-analysis

A network plot with pairwise comparisons of the relative performance of different concentration methods for concentration of enveloped and non-enveloped viruses based on their ranked performance in each published study is shown in Fig. 5. A total of 41 studies included enveloped viruses, and the plot shows that magnetic bead-based, precipitation and ultrafiltration are the most used and are usually preferred methods. In pairwise comparisons, most of the comparisons with (edges connected to) the magnetic bead-based method favour that method (4/5) and the average proportion of comparisons favouring that method is 63%, higher than for other methods (range 30-55%; captured by the node sizes in the plot). A total of 21 studies included non-enveloped viruses, and the plot shows that flocculation is on average the most preferred method (60% of pairwise comparisons favoured flocculation), with 3/5 edges favouring it (those from precipitation (8 studies), filtration and ultrafiltration).

**Fig. 5.**
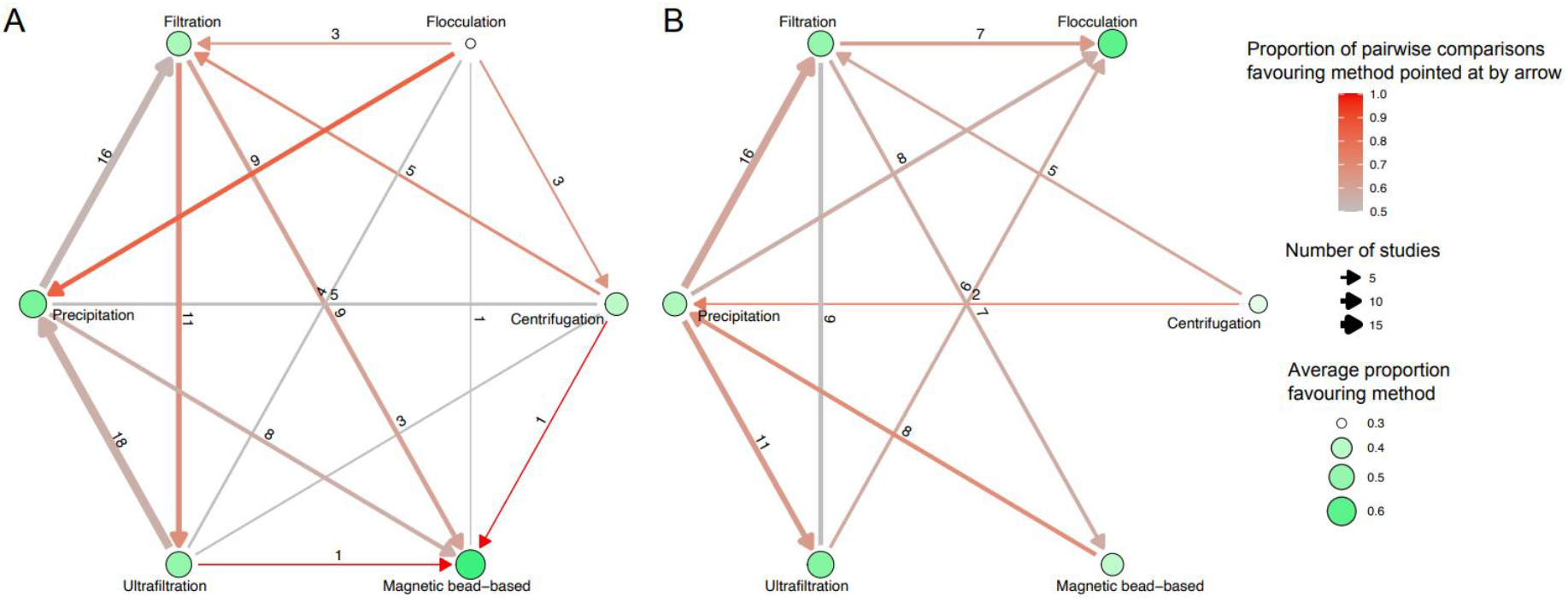
Network plots showing the pairwise comparison (arrows) between different concentration methods (nodes) for (A) enveloped viruses and (B) non-enveloped viruses. The arrows point to the method that was more sensitive in >=50% of comparisons and are labelled by the number of studies comparing those two methods. Arrow colour indicates the proportion of studies that favoured a given method based on their rank position in the studies. Node size and colour indicate the average proportion of comparisons that favoured that method. There is no arrowhead if across all pairwise comparisons in published studies the two methods were favoured equally. Nodes connected by edges represent the direct comparison (head-to-head studies comparing the connected concentration methods).

None of the methods for either enveloped or non-enveloped viruses were strongly favoured or disfavoured, with the proportion of studies favouring a particular method ranging between 30% and 63%.

In the Plackett-Luce model of the relative worth of different concentration methods, the magnetic bead-based method was ranked highest for enveloped viruses, followed by filtration, precipitation and ultrafiltration (Fig. 6). However, the estimated worth for each of the methods was not statistically significantly different compared to centrifugation (p-values > 0.05). In contrast to enveloped viruses, flocculation was shown to be the most effective method for non-enveloped viruses, followed by precipitation, magnetic bead based, ultrafiltration and filtration (again, all p-values > 0.05 compared with centrifugation; Fig. 6).

**Fig. 6.**
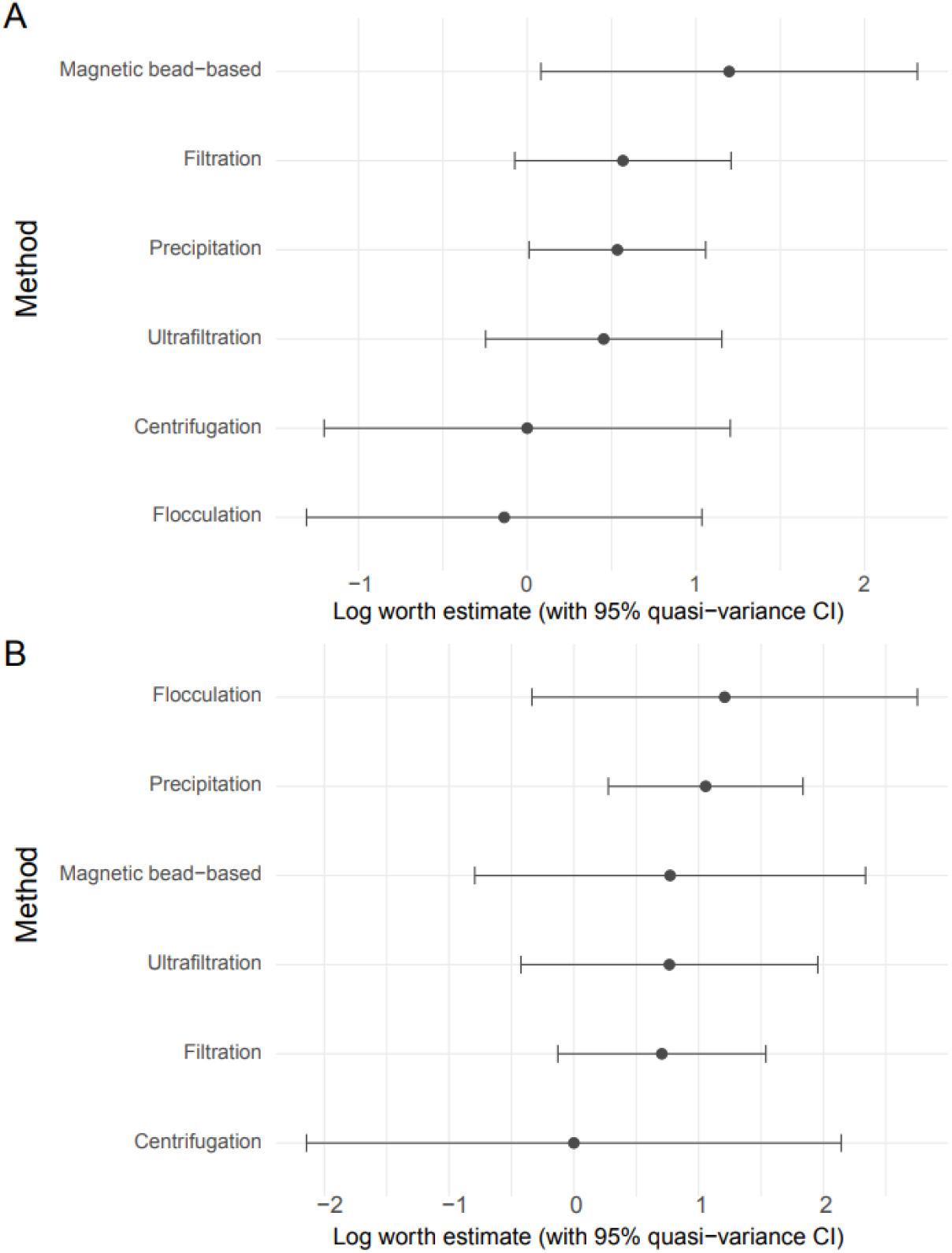
Estimated worth of each method based on Plackett-Luce analysis of their ranked performance across all studies. Estimated worth is shown on a log scale and is relative to centrifugation as a reference group and is shown in (A) for enveloped viruses and (B) for non-enveloped viruses. Approximate 95% confidence intervals are shown based on the quasi variance.

## Discussion and conclusion

The detection of infectious disease pathogens in wastewater has been practiced for nearly a century [42-44], and WES is now firmly established as a valuable tool for monitoring population-level infection dynamics [45, 46]. Central to the success of WES is the selection of an appropriate wastewater concentration method, as pathogen recovery efficiency is strongly influenced by the physicochemical properties of wastewater and the nature of the target. Wastewater physicochemical properties including turbidity, pH, organic matter, and solids content vary substantially depending on catchment population size, water usage patterns, and environmental conditions [47]. Consequently, a concentration method that performs well in one setting may be suboptimal in another, underscoring the need for context-specific method selection. There is no single concentration method that is universally optimal for all wastewater sample types and virus groups, and method selection must be informed by both wastewater characteristics and virus group.

To our knowledge, this study represents the first comprehensive systematic review and network meta-analysis evaluating the impact of wastewater concentration methods on the recovery of both enveloped and non-enveloped viral pathogens. Drawing on data from published studies investigating 36 viral targets, our analysis reveals clear virus group dependent trends in method performance. Specifically, the findings suggest that concentration methods effective for enveloped viruses may differ from those most suitable for non-enveloped viruses. These comparative insights provide a useful framework for prioritising methods based on pathogen characteristics and support more informed experimental planning in WES. Meta-analysis indicated that magnetic bead– based methods tended to perform best for enveloped viruses, whereas flocculation-based approaches showed higher relative performance for non-enveloped viruses. However, these trends did not reach statistical significance, reflecting substantial heterogeneity across studies and limitations in the available comparative data. Indeed, when summarising pairwise comparisons across studies, the rank for each method was highly variable and the proportion of studies favouring one method over another was often close to 50%.

The absence of a clearly superior method for either enveloped or non-enveloped viruses likely reflects multiple layers of methodological variability across the published studies. Concentration protocols used in the published studies differed not only in their principle (e.g., centrifugation, precipitation, flocculation, filtration, magnetic bead-based, or ultrafiltration) but also in the critical operational parameters such as starting sample volume, pre-treatment steps (e.g removal of solids or pH adjustment), elution buffers, incubation times, and centrifugation speeds and time. Even minor procedural differences can substantially affect how well the virus binds to filters (viral adsorption), partitions between solid and liquid fractions, and overall virus recovery efficiency, particularly for enveloped viruses such as coronavirus or influenza that have a more fragile outer layer that is sensitive to physicochemical disruption. If damage to the outer layer happens due to changes in temperature, chemicals, mixing, or handling, the virus may behave differently, making it harder to recover and detect accurately. In addition, wastewater samples used in each of the published studies likely varied widely in turbidity, organic load, suspended solids, and presence of inhibitors, all of which influence virus partitioning and recovery. However, most studies did not report on the wastewater properties, so it was not possible to include wastewater properties (e.g. pH, turbidity, and suspended solids) as a covariate in the assessment of method performance. Difference in whether viruses were endogenous or experimentally spiked, and in use of surrogate versus target pathogens, further complicated direct comparison.

Downstream detection platforms also introduce variability. Detection and quantification by qPCR, dPCR, NGS, or cell culture differs in analytical sensitivity, tolerance to inhibitors and ability to distinguish between infectious and non-infectious viral particles. For example, dPCR may provide greater precision at low concentration than qPCR [48], whereas cell culture assays assess infectivity rather than genome presence. The published studies also varied in how recovery efficiency was defined and calculated (e.g., percentage recovery of spiked/seeded virus, Ct value, limit of detection or detection rate).

Collectively, this heterogeneity across concentration, detection and analytical workflows likely attenuates observable performance differences between methods and limits the ability to identify an optimal approach for specific wastewater types and pathogen groups. Rather than indicating true equivalence, the absence of statistical superiority may therefore reflect insufficient harmonisation and standardised head-to-head evaluation across diverse wastewater matrices and virus types.

Despite considerable methodological advances, several challenges remain before wastewater concentration methods can be routinely applied in regulatory or decision-making contexts with high statistical confidence. For example, currently, only two concentration methods are formally standardised: the PEG-based two-phase separation method recommended by the World Health Organization for poliovirus surveillance and the virus adsorption–elution (VIRADEL) method described in US EPA Method 1615. In addition, depending on the approach used, pathogen loss during processing or co-concentration of PCR inhibitory substances may occur. Wastewater contains substantial amounts of organic matter, humic and fulvic acid and tannins, as well as organic substances resulting from human activity (e.g detergents, pesticides, hydrocarbons and pharmaceuticals). When samples are concentrated to detect low-abundance pathogens, these substances are often concentrated concurrently, potentially inhibiting downstream molecular or cell culture-based assays. Finally, the inherent heterogeneity of wastewater composition presents a major obstacle to the optimisation and standardisation of concentration methods.

In conclusion, wastewater concentration methods exert significant influence on the overall success of pathogen detection by shaping the efficiency of downstream extraction and detection methods. No single method is universally applicable across all wastewater matrices or virus types. Instead, method selection should be guided by the characteristics of the wastewater sample, the pathogen(s) of interest, and the specific objectives of surveillance. Continued efforts toward protocol standardisation, coupled with targeted comparative studies, will be essential to strengthen the reliability, comparability, and public health utility of wastewater-based infectious disease surveillance.

## Methods

### Search strategy and study selection

Two databases, PubMed, and Web of Science, were used for studies on the effectiveness of wastewater concentration methods in virus recovery and capture from wastewater and/or sewage samples. All retrieval results from the database were completed on January 31^st^, 2025. The search strategy in PubMed included the following keywords: ((((wastewater) AND (sewage)) AND (concentration method)) AND (concentration methods)) AND (wastewater surveillance).. The Web of Science search strategy included the keywords: (“wastewater” OR “wastewater surveillance” (All Fields)) AND “concentration methods” (All Fields).

### Eligibility criteria

The titles and abstracts of all identified articles were assessed for eligibility based on the following inclusion criteria: (1) studies comparing two or more concentration methods for wastewater or sewage, specifically for detecting viral pathogens; (2) quantitative studies with conclusive result reporting pathogen recovery efficiency, statistical results, and detection limits. Exclusion criteria were as follows: (1) systematic reviews, conference papers, protocols; (2) studies not comparing or conducting a head-to-head evaluation of the performance of wastewater concentration methods; (3) studies comparing methods for collecting microplastics, pharmaceutical compounds, organic pollutants in wastewater; (4) studies focusing on wastewater treatment; (5) studies comparing variants of one group/type of concentration method; (6) wastewater surveillance studies with no wastewater concentration methods. The relevant literature was independently searched and screened by two authors (Akello and Bellekom). R environment software was utilized to delete duplicates.

### Data extraction and ranking

A data extraction document was created to extract suitable data including (1) study characteristics (authors, title, and publication year); (2) pathogen characteristics (enveloped and/or non-enveloped viruses, virus species); (3) details of wastewater concentration analysis (methods evaluated, viral pathogens assessed); (5) outcome measures (results of best performing methods based on virus recovery efficiency, mean Ct values, yielded a greater number of positive samples, and lower limit of detection of assay). Data extraction was independently conducted by two authors (Akello, and Bellekom), with disagreements resolved through discussion with the other authors (Shaw, and Grassly). Concentration methods were classified by their main concentration principle (1. Centrifugation, 2. Filtration, 3. Flocculation, 4. Magnetic bead, 5. Precipitation, 6. Ultrafiltration). Within each study, concentration methods were ranked based on reported virus recovery efficiency, defined primarily as the proportion of spiked or seeded virus successfully recovered following concentration and quantified by downstream molecular assays including qPCR, ddPCR and dPCR. Where direct recovery percentages were no reported, related performance outcomes were used, including Ct values, higher detection rates across replicate samples, and assay limits of detection (indicating improved analytical sensitivity). Together, these outcome measures were used as comparative proxies of concentration performance within each study to enable relative ranking across methods. Ties were permitted where studies reported no statistically significant difference between methods. Virus recovery efficiency was considered the primary indicator of concentration method performance.

### Data analysis

The performance of different concentration methods was visualised in a network plot, where nodes corresponded to each method and edges were labelled according to the proportion of times one method ranked above the other in any comparisons that included those two methods. Overall performance of each method for enveloped and non-enveloped viruses was analysed using the Plackett–Luce ranking model implemented in R [49, 50]. This probabilistic model estimates a “worth” parameter for each method, representing the probability that a given concentration approach outperforms others in recovering viral targets from wastewater samples in any given comparison experiment. The model was applied to ranked data comprising six major concentration method categories with tied rankings allowed when pairwise comparisons were reported as not statistically significantly different.

## Data Availability

All the data produced in the present work are contained in the manuscript or it's supplementary materials

## Supplementary Material

*Supplementary file 1*. Detailed narrative reviews of each concentration method, including methodological variants and performance considerations.

*Supplementary file 2*: Excel spreadsheet file with the information of the studies included.

## Acknowledgements

We thank members of the Vaccine Epidemiology Research Group for useful discussions. This study was supported by the Gates Foundation under grant agreement INV-076271. All authors acknowledge funding from the Medical Research Council (MRC) Centre for Global Infectious Disease Analysis (reference MR/X020258/1), funded by the UK Medical Research Council. This UK-funded award is carried out in the frame of the Global Health EDCTP3 Joint Undertaking.

