## Supplementary File 1 for "Principles and performance of wastewater concentration methods for environmental surveillance of viruses: a systematic review and meta-analysis"

1. **Wastewater concentration – A critical step in Implementing WES for pathogen detection**

Wastewater is a complex matrix composed of approximately 99.9% water and 0.1% dissolved and suspended constituents, including pathogens (such as viruses, bacteria, fungi, protozoa, and helminths), nutrients, solids, and a wide range of organic and inorganic compounds (Fig. 1). Concentration methods for capture and recovery of pathogen particles and/or their genomes play a critical role in the successful implementation of WES, as they enable reliable detection and characterisation of pathogens present at low abundance and support public health responses. Based on their underlying principles /mechanisms, wastewater concentration methods can be broadly categorised into six groups: centrifugation, filtration, flocculation, magnetic bead-based approaches, precipitation, and ultrafiltration (Fig. 1). To date, most concentration methods have been optimised for specific pathogens or for limited groups of related organisms, such as enteric viruses. Although considerable efforts have been made to find a single concentration method capable of simultaneously recovering multiple viral, bacterial, and fungal pathogens from wastewater, these attempts have so far yielded mixed results. Consequently, no universal concentration method currently exists that is suitable for all pathogens. This limitation reflects both the inherent complexity of the wastewater matrix and the multiple physical and chemical steps involved in concentration procedures, which can interfere with downstream detection for example, through the co-concentration of PCR inhibitors. Several studies have demonstrated that the performance of wastewater concentration methods varies according to wastewater sample characteristics and pathogen properties; for viruses, in particular, differences in recovery efficiency have been observed between enveloped and non-enveloped viruses.

1.1. **Centrifugation based methods**

Centrifugation is a commonly used first step prior to pathogen concentration in wastewater samples particularly before ultrafiltration as solid matter may cause membrane clogging. Methods eliminating solid matter in the first step by centrifugation includes ultracentrifugation and ultrafiltration. Centrifugation and ultracentrifugation work based on the sedimentation principle, which states that the denser particles settle down faster when compared to less dense particles under gravity. During ultracentrifugation, the sample is spun around an axis, generating a centrifugal force that acts on various particles within the sample. Larger molecules move quickly, while smaller molecules move slowly. Simultaneously, denser molecules are pushed outward to the periphery of the tubes, while less dense molecules are drawn toward the centre of the tube. Once the process is completed, the larger and more dense particles settle down, forming pellets at the bottom of the tube. In comparison, the smaller and less dense particles remain either suspended in the supernatant or float on the surface. Ultracentrifugation has been implemented for routine WES in Hong Kong [1]. Although ultracentrifugation is appealing due to its high pathogen recovery efficiencies, it is not readily available in many laboratories as it relies on the expensive equipment e.g the Beckman Coulter ultracentrifuge costs between $300,000 to $500,000.

1.2. **Filtration-based methods**.

Filtration-based methods using electronegative or electropositive filters are the most widely used methods for the primary concentration of wastewater [2, 3]. These methods have been widely used to concentrate enteric viruses from both untreated and treated wastewater. Their development is based on electrostatic interactions between viruses and filter materials, utilising the fact that most enteric viruses carry a net negative charge in environmental waters at near-neutral pH [4-7]. In this method, negatively charged viral particles adsorb directly onto electropositive filters or onto electronegative filters through salt bridging mediated by multivalent cations. Following adsorption, viruses are typically eluted from filters using approximately 1–1.6 L of elution buffer [8, 9]. Several procedures of eluting absorbed viruses from filters have been described, including the use of different buffers such as beef extract, glycine buffer, and urea–arginine phosphate buffer, and use of either single or multiple elution steps [10-12]. Karim *et al.* (2009) demonstrated that a two-step elution protocol yielded higher viral recoveries than a single elution, with optimal performance achieved by immersing the filter in beef extract for 1 min during the first elution and 15 min during the second. In contrast, extending the second elution overnight resulted in reduced viral recovery, which the authors attributed to viral inactivation caused by prolonged exposure to alkaline pH (pH 9) and incubation at room temperature [8].

Comparative studies have shown that electronegative and electropositive filters, as well as ultrafilters can perform similarly for viral concentration from wastewater, with differences in recovery largely driven by virus type rather than filter type, wastewater matrix, or sample volume [2]. However, unlike electropositive filtration, electronegative filtration requires preconditioning of wastewater samples, making it less applicable across a wide range of wastewater quality conditions [3]. Filtration-based methods are also limited to low-turbidity samples, as high particulate loads can cause filter clogging. In addition, their ability to recover multiple pathogen types simultaneously (viruses, bacteria, and protozoa) is limited, owing to the need for pathogen-specific elution protocols. Substantial variability in recovery efficiency among different enteric viruses has been reported [13].

Various electropositive microfilters composed of different materials is commercially available, including NanoCeram filters (Argonide, USA), Zeta Plus 50S and 60S disc filters (3M Purifcation Inc. USA), Seitz filters (Pall Corporation, Germany), and 1MDS filters (3M Purifcation Inc. USA). While these filters are designed primarily to capture viruses via electrostatic attraction, pathogens may also attach to the filter through hydrophobic interactions or size exclusion [14]. Among these, the 1MDS electropositive microfilter is the most widely used and is recommended by the United States Environmental Protection Agency (US EPA) for concentrating human enteric viruses from water samples using the virus adsorption–elution (VIRADEL) method. Although 1MDS filters effectively retain viruses, bacteria, and protozoa, elution and recovery efficiencies vary across organism types, and the filters are not cost-effective for routine viral monitoring [8, 14]. Karim *et al.* (2009) reported that NanoCeram filters achieved viral recoveries comparable to those of 1MDS filters, supporting their use as a lower-cost alternative [8]. Positively charged NanoCeram filters have since been incorporated into the US EPA’s proposed Method 1615. In China, filtration using mixed cellulose ester membranes has been adopted for routine environmental surveillance of enteroviruses since 2008 [15-17]. A comparative study by Fang *et al.* demonstrated that this filtration-based approach exhibited higher sensitivity for enterovirus detection in sewage than the conventional two-phase concentration method [17].

1.3. **Flocculation-based methods**

Flocculation-based methods rely on the adsorption of pathogens onto floc aggregates, enabling their subsequent recovery from wastewater. Aluminium-based flocculation has been shown to achieve a lower limit of detection for SARS-CoV-2 (4.3 × 10² genome copies (GC)/mL) compared with PEG precipitation (4.3 × 10³ GC/mL), indicating superior analytical sensitivity [18]. Banadaki *et al.* reported high recovery efficiencies (41%) for SARS-CoV-2 using the calcium flocculation–citrate dissolution (CFCD) method, which offers rapid processing (<45 minutes) and low operational costs (<US$2 for four sample replicates) [19].

Skimmed-milk flocculation represents a particularly accessible approach, as it does not require specialised laboratory infrastructure and relies on readily available consumables. These characteristics make it a promising option for wastewater and environmental surveillance in resource-limited settings, where affordability and supply chain reliability are critical for sustaining uninterrupted surveillance [20].

1.4. **Magnetic bead-based method**

*Nanotrap Particles*

The Nanotrap Particles developed by Ceres Nanoscience, Manassas, VA, are hydrogel polymer particles consisting of allylamine, N-isopropylacrylamide, and crosslinked with N, N’-methylenbisacrylamide. Nanotrap Particles are functionalised with various dye affinity baits that facilitate capture, binding and enrichment of low abundance pathogens from complex biological matrixes including wastewater and concentrate them into smaller volumes [21]. As Nanotrap Particles may be of negative or positive charge, they can attract and capture viruses through the interaction of the Nanotrap Particle with the negative or positively charged residues on the surface of viral pathogens. Although they were originally designed to harvest proteins and peptides, recent findings by Shafagati *et al.* demonstrated the utility of Nanotrap particles in capturing and detecting virions [21]. The Nanotrap Microbiome A and B particles can capture and concentrate multiple pathogens including viruses , bacteria, and protozoan parasites in one sample [22, 23]. Since the COVID-19 pandemic, Nanotrap particles have been utilised to detect SARS-CoV-2 and other respiratory viruses in wastewater [19, 23-25]. Like many other concentration methods, the Nanotrap Particles may recover certain pathogens better than others [22]. Employing automated Nanotrap workflows for WES can reduce turnaround time. Karthikeyan *et al*. showed that the turnaround time was reduced by 20-fold following processing of samples by automation compared to manual (5 hours for 96 samples from processing to result). Additionally, this study found that miniaturization of the sample processing pipeline significantly reduced the processing cost to $13/sample [25] . Liu *et al.* showed that the automated Nanotrap workflow had significantly higher SARS-CoV-2 recoveries compared to the membrane filtration and the Skimmed-milk flocculation [26]. In this study, the authors used different extraction kits for the Nanotrap workflow and membrane filtration workflow which could have impacted the results. The manual Nanotrap workflow can be used in resource-limited settings.

*The porcine gastric mucin coated magnetic bead (PGM-MB)*

The porcine gastric mucin coated magnetic bead (PGM-MB) based method is such a promising technique that can be utilised to simply concentrate, capture, and recover multiple viruses from wastewater. PGM is a biological substrate comprised essentially of glycoproteins such as glycans that act as virus receptors. The principle of the PGM method is based on virus binding to the PGM glycans which is thought to mimic host receptor binding step in viral infection. The ability of viruses to selectively bind to PGM has been reported [27] and has been used for the recovery of viruses from complex matrixes including sewage, stool, and food samples, as well as to differentiate between infectious and non-infectious virions [28-31]. Oh *et al*. in their study demonstrated that human and animal viruses were efficiently concentrated from wastewater using PGM-MB [28]. This finding demonstrates the possibility of utilizing PGM-MB as a cost-effective viral concentration method for direct detection of emergent zoonotic pathogens. In this respect, virus concentration using PGM-MB may be a promising and cost-effective technique that can be utilised to quickly and simply, concentrate, capture, and recover emergent viruses from sewage.

1.5. **Precipitation-based methods**

Precipitation-based methods are a cornerstone of pathogen recovery from wastewater, allowing the separation and concentration of pathogens from wastewater by altering the solubility of the sample matrix. These approaches typically involve the addition of precipitation reagents such as, salts, solvents or polymers, often combined with pH adjustment to alter solubility of the sample. This causes pathogens and other macromolecules to settle out of the water as a solid precipitate. The resulting precipitate can then be recovered by centrifugation or filtration, while the remaining supernatant (liquid) often containing smaller pathogens that do not precipitate out of the sample may also be analysed. Precipitation methods can be applied either as a primary concentration step during initially sample processing or as a secondary concentration method following pathogen elution from filters or membranes.

Common precipitants include aluminium chloride (AlCl_3_), polyethylene glycol (PEG), and magnesium chloride (MgCl_2_). Although no significant differences in overall viral recovery efficiency have been consistently observed among these precipitants, AlCl₃ precipitation has been reported to exhibit lower variability across samples [1]. PEG-based protocols, including approaches that either remove solids or retain solids, are among the most widely used precipitation methods and have been applied predominantly for viral concentration, particularly for non-enveloped viruses. PEG is a hydrophilic polymer that can induce the precipitation of pathogens (viruses and bacteria), and other macromolecules by excluding water molecules and reducing solubility.

The PEG/dextran method, also known as the two-phase method, relies on liquid–liquid partitioning to separate pathogens based on their physicochemical properties [32, 33]. In this process, wastewater samples are first clarified by centrifugation to pellet large, suspended solids, which are processed separately since pathogens may be partially bound to these solids. The clarified supernatant is then mixed with PEG and dextran and incubated at low temperature (4° C) to allow phase separation. This process results in two immiscible aqueous phases: an upper PEG-rich, relatively hydrophobic phase and a lower dextran-rich, denser, more hydrophilic phase. Under appropriate salt and pH conditions, most pathogens partition into the lower phase or accumulate at the interface. The separation occurs in a funnel, allowing the lower phase and interface to be collected without disturbing the upper phase. The pellet from the initial centrifugation is then added to this concentrate. The two-phase method is currently recommended by the World Health Organization for poliovirus concentration in environmental surveillance.

Although Fang *et al.* reported that the two-phase method was less sensitive than filtration using mixed cellulose ester membranes for enterovirus detection in sewage, it has nevertheless been extensively and successfully used in poliovirus environmental surveillance programmes worldwide [17, 34-37]. Its long-standing implementation has contributed substantially to the detection and tracking of poliovirus circulation.

The performance of PEG precipitation is strongly influenced by operational parameters. Increasing PEG molecular weight has been shown to enhance sensitivity, whereas increasing centrifugal force during PEG precipitation does not significantly improve viral recovery [38]. Considerable variability in reported recovery efficiencies has been observed across studies, reflecting differences in protocol design, surrogate viruses, wastewater characteristics, and analytical workflows. Torri *et al.* compared five PEG-based methods with varying operational parameters for the recovery of murine hepatitis virus (MHV), bacteriophage φ6, pepper mild mottle virus (PMMoV), and murine norovirus from 34 raw wastewater samples collected in Japan. Recovery efficiencies varied widely, ranging from 0.07% to 2.6% for MHV and from 7.6% to 89% for other targets [39]. PEG precipitation with a 2-h incubation consistently outperformed protocols employing overnight incubation, although Flood *et al.* reported no statistically significant difference between PEG procedures with and without overnight incubation [40]. Together, these studies indicate that PEG-based methods can be adapted to support same-day processing without substantial loss of sensitivity.

Further variability in PEG performance, particularly for enveloped viruses, has been attributed to differences in incubation time, sample pretreatment, surrogate selection, wastewater quality, and biases in quantifying seeded virus loads[39]. Wu *et al.* refined a PEG-based protocol for SARS-CoV-2 concentration and reported the most consistent recoveries from pellets obtained using 0.2-µm filtrates [41]. Despite their broad applicability, PEG precipitation methods generally show lower recovery efficiencies for enveloped viruses than for non-enveloped viruses, likely because enveloped viruses preferentially associate with wastewater solids [42]. This limitation has been corroborated by multiple studies, including those by Ahmed *et al.*, which identified PEG precipitation as one of the least effective approaches for enveloped virus recovery [43].

Precipitation-based methods remain attractive due to their simplicity, low cost (for example, <US$2 per sample for PEG-based approaches compared with >US$34 for ultrafiltration methods), and compatibility with a wide range of wastewater volumes and turbidity levels [40, 44, 45]. Additionally, pathogen precipitation under low-temperature, high-salt conditions can stabilise viral particles and provide an isotonic carrier medium conducive to downstream analysis [44]. Although PEG protocols are often considered labour-intensive because of long incubation and centrifugation steps, optimisation studies have demonstrated that processing time can be reduced without significantly compromising sensitivity, thereby increasing testing throughput [46].

1.6. **Ultrafiltration-based methods**

Ultrafiltration (UF) is a size-exclusion–based concentration approach that separates and retains microorganisms using semi-permeable membranes rather than electrostatic interactions. As a result, UF enables the simultaneous concentration of a broad range of pathogens including viruses, bacteria, fungi, protozoa, and parasites based on particle size. UF is probably the most widely used method for concentrating SARS-CoV-2 particles and viral genomes from wastewater. In India and Bangladesh, both Moore swab and ultrafiltration approaches have been successfully applied to concentrate *Salmonella enterica* serovar Typhi and *Salmonella enterica* serovar Paratyphi A from wastewater samples [47].

Commercial UF devices such as Amicon and Centricon (Merck Millipore, Germany) employ membranes defined by molecular weight cut-off rather than pore size, making them attractive for multi-pathogen recovery. However, their performance can be compromised by the high organic matter typically present in wastewater, which may cause the membrane to collapse and limit the volume that can be processed often to approximately 100 mL. Consequently, UF methods are less suitable for field applications or highly turbid samples. Following concentration, viruses retained on UF membranes are typically eluted using organic (for example, beef extract) or inorganic (such as sodium polyphosphates) solutions. The resulting eluates may be subjected to a secondary reconcentration step to further reduce volume and enhance sensitivity for downstream detection methods (molecular assays or cell-culture–based infectivity assays). Although UF membranes generally exhibit high virus retention efficiency, elution and reconcentration success varies considerably, reflecting the biological diversity of viruses and the complexity of wastewater matrices.

Amicon ultrafiltration devices are most suitable for low-volume samples (≤20 mL). Processing larger volumes requires pre-filtration through 0.45-µm and/or 0.2-µm filters to remove debris. While Amicon filtration has been reported to yield relatively high viral recoveries, substantial variability has been observed across virus types and between replicates [45]. In addition, the final concentrate volume may be inconsistent and, in some cases, too high for certain nucleic acid extraction protocols. Ultrafiltration processing times are also highly dependent on wastewater characteristics, with prolonged centrifugation potentially affecting pathogen integrity and recovery [45]. Notably, Farkas *et al.* reported that Amicon ultrafiltration may perform particularly well in catchments with elevated ammonium concentrations, such as areas influenced by agricultural runoff [45]. An optimised UF protocol incorporating a 0.22-µm pre-filtration step has also been shown to improve recovery of enveloped viruses [42].

InnovaPrep ultrafiltration systems enable rapid concentration of pathogens from wastewater volumes of up to 50 mL, although reported viral recoveries are generally lower than those obtained using Amicon devices. However, the incorporation of sonication during ultrafiltration, as implemented in the InnovaPrep Concentrating Pipette Select system, has been shown to enhance virus recovery [48].

Tangential flow hollow-fiber ultrafiltration (TFUF), also known as crossflow filtration, represents an alternative UF method in which the sample flows tangentially across the membrane surface. This design reduces membrane clogging by preventing particle accumulation, while applied pressure allows smaller molecules to pass through the membrane. Larger components are retained and recirculated for further concentration. TFUF is typically used as a primary concentration step and is well suited for processing large wastewater volumes (>1 L). Using hollow-fibre membranes with a molecular weight cut-off of approximately 100 kDa, TFUF has been shown to efficiently recover a range of microorganisms [2, 3, 49-51]. The ability of TFUF to co-concentrate multiple pathogens makes it particularly valuable for comprehensive assessment of microbial water quality.

2. **Consideration for wastewater concentration method selection**

Despite substantial progress in the development and optimisation of wastewater concentration methods, selecting an appropriate concentration method for WES requires careful consideration of multiple factors, including sample type, volume, composition, and pathogen-specific characteristics. These variables can substantially influence concentration method performance in capturing and recovering pathogens for downstream. Farkas *et al.* demonstrated that, irrespective of the concentration method employed, viral recovery from wastewater is strongly affected by sample volume and matrix composition, as well as by viral characteristics [45]. This consideration is particularly important given the widespread reliance on PCR-based detection methods, which are sensitive to both low target abundance and the presence of inhibitory substances. Accordingly, researchers implementing WES should systematically evaluate the key factors that influence the performance of wastewater concentration methods prior to method selection (Fig. S1), to ensure reliable and interpretable surveillance outcomes.

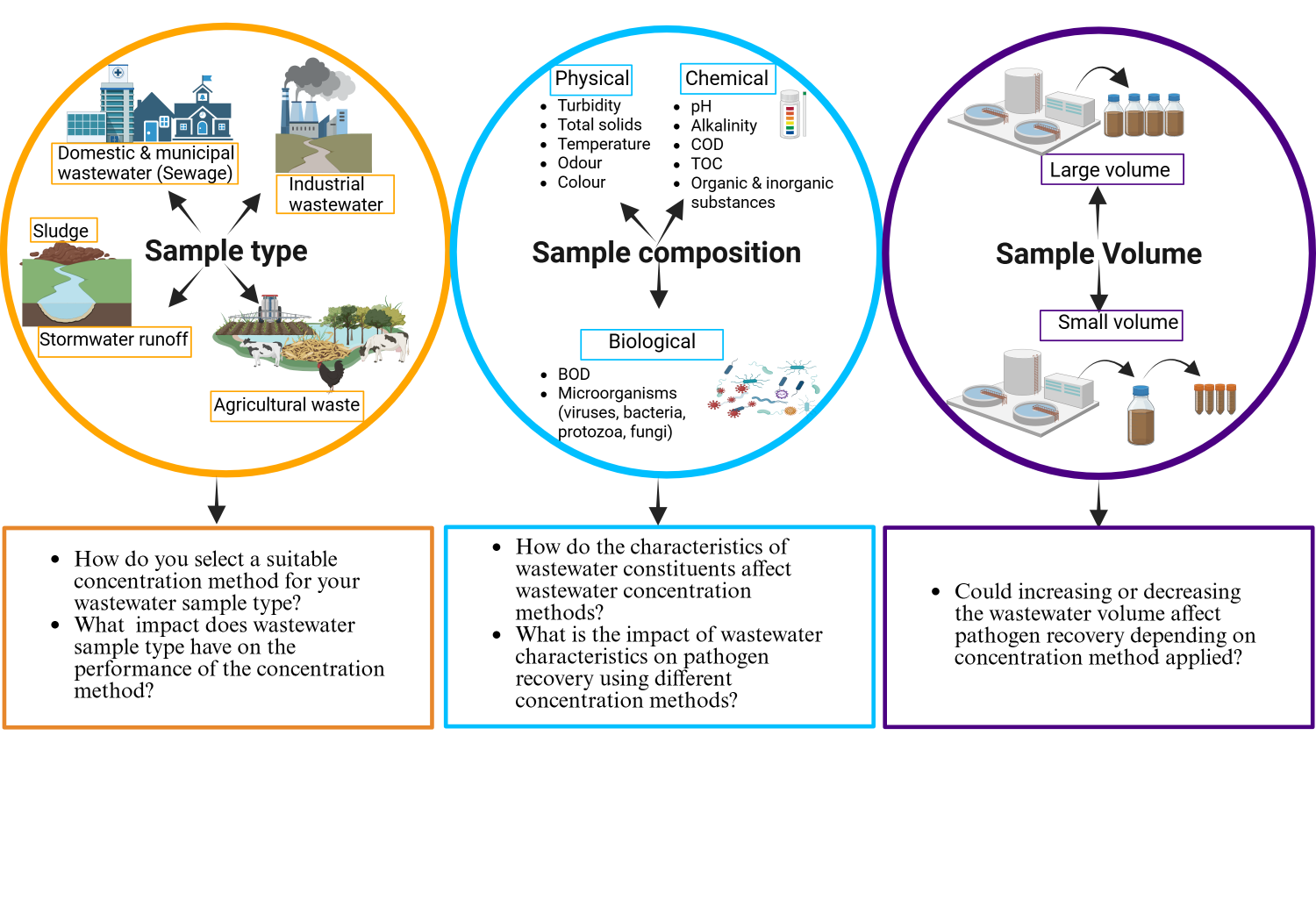

**Fig. S1.** Overview of the three key factors influencing wastewater pathogen concentration performance. Variability across these domains can influence viral partitioning, adsorption of viruses to filters, co-concentration of PCR inhibitors, and downstream detection efficiency. The figure highlights the multifactorial nature of wastewater matrices and the importance of selecting and optimising the concentration method based on these three key factors. Created in BioRender. <https://BioRender.com/xvqu6su>.

2.1. **Sample type**

The choice of wastewater concentration method can be influenced by sample type, as pathogen abundance, solids content, and inhibitory substances vary substantially between wastewater matrices. Filtration and precipitation-based methods are generally well suited for untreated wastewater (influent), whereas centrifugation-based methods are most effective for primary sludge, where pathogens are predominantly associated with settled solids. Raw untreated wastewater is commonly used for surveillance of enteric viruses and SARS-CoV-2 but typically requires the processing of large volumes (ranging from approximately 500 mL to >1,000 mL) due to extensive pathogen dilution. Suitable concentration methods for influent samples include precipitation-based methods and electronegative membrane filtration using the virus adsorption–elution (VIRADEL) method. However, membrane-based filtration methods are prone to clogging in high-turbidity wastewater. Ultrafiltration systems, such as Amicon or Centricon devices, are better suited for low-turbidity influent samples and generally process smaller volumes (approximately 10–50 mL). Skimmed-milk flocculation also represents a viable option for raw wastewater, particularly in resource-limited settings. In some workflows, centrifugation or ultracentrifugation is used as an initial clarification step to remove large particulates prior to applying downstream concentration methods.

In contrast, primary sludge consists largely of settled solids and contains higher concentrations of pathogens, allowing for smaller input volumes (typically ~100 mL) to be analysed. Concentration methods commonly applied to primary sludge include centrifugation and ultracentrifugation, which effectively recover pathogens bound to solids. Magnetic bead–based methods, such as Nanotrap particles (Ceres Nanosciences, USA), have also been successfully used for both raw wastewater and primary sludge samples. Notably, SARS-CoV-2 RNA is often detected at higher concentrations in primary sludge than in raw influent wastewater. However, concentrating pathogens from primary sludge presents additional challenges, as this matrix contains high levels of PCR inhibitors that are not always fully removed during concentration and nucleic acid extraction. These inhibitors can interfere with downstream molecular analyses, necessitating careful optimisation of both concentration and purification steps.

2.2. **Sample volume**

Early wastewater concentration techniques developed in the 1960s and 1970s focused on recovery of enteric viruses from small sample volumes. To date, most published virus concentration protocols process between 30 and 250 mL of wastewater, reflecting practical constraints related to sample handling in the laboratory and the availability of specialised equipment. Although some studies have proposed that increasing wastewater sample volume and concentration factor may enhance detection sensitivity [52], empirical evidence supporting this approach remains inconsistent. For example, Zhou *et al.* reported improved poliovirus detection using a bag-mediated filtration system (BMFS) compared with the two-phase method, an effect attributed to the substantially larger volume processed (1,620 mL versus 150 mL) [53]. In contrast, Farkas *et al.* observed that increasing wastewater volume negatively affected viral recovery efficiency, while Zheng *et al.* found no significant improvement in sensitivity associated with large-volume sampling [1, 45].

Further comparisons indicated that although ultracentrifugation applied to large volumes outperformed AlCl₃ precipitation and membrane adsorption methods processing similarly large volumes, its sensitivity was comparable to that achieved using ultracentrifugation with small sample volumes (30 mL). Based on these findings, the authors concluded that ultracentrifugation using smaller input volumes is preferable due to higher sample processing throughput [1]. Taken together, these studies indicate that increasing wastewater sample volume does not consistently improve viral detection sensitivity and may, in some cases, reduce recovery efficiency. This indicates that optimisation of concentration protocols and overall processing efficiency appears to be more critical than sample volume alone for effective wastewater surveillance.

2.3. **Sample composition**

Wastewater composition also referred to as wastewater characteristics includes the physical, chemical, and biological constituents present in wastewater. The wastewater composition/ characteristics are influenced by its source and can vary significantly depending on the local conditions. The physico-chemical characteristics of wastewater can exert significant influence on the performance of the concentration method affecting capture and recovery of pathogen particles and/or their genomes. Drawing on published studies and research on efficiency of different concentration methods for WES, understanding wastewater characteristics is crucial for selecting the appropriate concentration methods for the specific research objective, as they can have a considerable impact on performance of the concentration method used.

*Physical characteristics of wastewater*.

Physical characteristics such as turbidity, total solids (suspended and dissolved solids), and the temperature of wastewater are the main parameters characterising the impact of wastewater physical characteristics on the performance of the concentration methods. Turbidity which is the presence of suspended solids and colloidal particles, making the water appear cloudy is highly variable within and between wastewater samples [22]. Ahmed *et al.* showed that PMMoV recovery proportions by the Nanotrap Particles decreased with increasing turbidity whlist PMMoV recoveries by absorption extraction were not affected by either turbidity or total solids levels[22]. Wastewater samples with high levels of suspended solids such as organic matter and/or with high conductivity tend to have pathogen particles attached to the solid matter. Therefore, concentration methods eliminating solid matter can result in low pathogen titres. Suspended solid or soluble organics have been shown to affect adsorption of some viruses to electronegative filters. Studies by Farkas *et al.* demonstrated that only a small proportion of viruses attach to the solid particles in the pellet fraction[45] whereas other studies found that a relatively large proportion of viruses can be recovered from the pellet fraction[42]. The conflicting findings imply that there may be substantial differences in the ability of viruses/pathogens to adsorb to solid matter in different wastewater samples. Kitamura *et al.* showed that recovery of SARS-CoV-2 from the solid fraction rather than the supernatant fraction of wastewater is a more effective method [54].

Wastewater temperatures vary based on the source and geographic location. Increased temperature has been shown to accelerate the denaturation of viral nucleic acids and proteins by enhancing extracellular enzyme activity[55]. The effect of temperature on virus inactivation is thought to be impacted by various factors including organic content and the presence of antagonistic microorganisms present in wastewater. Low temperatures are required for concentrating enveloped viruses. Kevill *et al.* showed that sample turbidity, storage temperature, and surfactant load affected viral recovery, highlighting the need for careful consideration of the concentration method used when working with wastewater samples[56]. Sample turbidity has been reported to affect virus recovery from wastewater samples. In environments with extremely high wastewater turbidity and surfactant load, it is recommended to use a precipitation method rather than an ultrafiltration method. It is therefore important for researchers conducting WES for pathogen detection to measure the turbidity and total suspended solids of wastewater samples that they are conducting analysis on to ensure that they use a suitable concentration method.

*Chemical characteristics of wastewater*

Chemical characteristics important for pathogen concentration in wastewater include pH, alkalinity, chemical oxygen demand (COD), total organic carbon (TOC), organic and inorganic substances, heavy metals, and trace elements. These characteristics impact the efficiencies of concentration methods as different contaminants may affect the various concentration methods differently, with some being more or less affected than others. Viruses once released into the environment are susceptible to inactivation due to virucidal effects caused by factors such as temperature, pH, UV light, inorganic cations and anions, and antagonistic microbial interaction. Neutral pH is required for concentrating enveloped viruses. Precipitation-based methods are robust and more resilient to organic matter than ultrafiltration [57]

*Biological characteristics of wastewater*

Biological characteristics of wastewater include biochemical oxygen demand (BOD) and microorganisms that are due to contaminants. Pathogens particularly viruses, differ drastically in structure, genetic variability, chemical and mechanical resistivity. This diversity contributes to varied efficacy and sensitivity to the methods that greatly influence viral detection. Enteric and respiratory pathogens are usually stable in wastewater. Non-enveloped enteric viruses are known to be more resistant to environmental factors and often survive longer than enveloped viruses[58]. Enteric viruses such as enteroviruses, norovirus, adenovirus, astrovirus, hepatitis A and E virus, and rotaviruses are often detected in wastewater because they are shed in high concentrations in the stool of symptomatic and asymptomatic individuals. Arboviral concentration in wastewater hasbeen suggested to be lower compared to other viruses.

Inactivation of coronaviruses in water has been reported to be dependent on temperature, level of organic matter and presence of antagonistic bacteria. Wastewater concentration methods may vary in efficiency based on the pathogen characteristics including structure and size [59]. Enveloped viruses tend to be sensitive to some organic solvents used in concentration methods. Some concentration methods such as precipitation and flocculation methods that are commonly used to concentrate non-enveloped viruses from wastewater samples may be unsuitable for enveloped viruses due to these viruses being relatively unstable in the environment and more susceptible to common oxidants [60]. Therefore, concentration methods may need to be selected or adapted for different viruses depending on their structure, morphology and physical properties.

Moreover, most wastewater concentration methods have been developed and tailored to recover enteric pathogens, particularly non-enveloped viruses such as polioviruses, noroviruses, adenoviruses, enteroviruses, and hepatitis A virus that tend to be found in the supernatant fraction, and they have been shown to be inefficient to recover enveloped viruses such as SARS-CoV-2 from wastewater [42, 61]. Poor adsorption of non-enveloped enteric viruses to wastewater solids has been reported [62, 63]. Despite this, some enteric viruses have been observed in primary settled solids in high concentrations [64, 65]. Of the eleven concentration methods evaluated by Barril *et al.* 2021 for SARS-CoV-2 recovery, PEG precipitation and aluminium flocculation had the highest SARS-CoV-2 recoveries from wastewater [18].
